# Increased cortical excitability and reduced brain response propagation during attentional lapses

**DOI:** 10.1101/2020.04.01.20049650

**Authors:** Paolo Cardone, Maxime Van Egroo, Daphne Chylinski, Justinas Narbutas, Giulia Gaggioni, Gilles Vandewalle

## Abstract

Modern lifestyle curtails sleep and increases nighttime work and leisure activities. This has a deleterious impact on vigilance and attention, exacerbating chances of committing attentional lapses, with potential dramatic outcomes. A full characterization of the brain mechanisms associated with lapses is still lacking. Here, we investigated the brain signature of attentional lapses and assessed whether cortical excitability and brain response propagation were modified during lapses and whether these modifications changed with aging. We compared electroencephalogram (EEG) responses to transcranial magnetic stimulation (TMS) during lapse and no-lapse periods while performing a continuous attentional/vigilance task at night, after usual bedtime. Data were collected in healthy younger (N=12; 18-30 y) and older individuals (N=12; 50-70 y) of both sexes. Amplitude and slope of the first component of the TMS-Evoked Potential (TEP) and Response Scattering (ReSc) were used to assess cortical excitability and brain response propagation, respectively. In line with our predictions, TEP during lapses was characterized by larger amplitude and slope. We further found that ReSc over the cortical surface was lower during lapses. Importantly, cortical excitability increase and response propagation decrease during lapse did not significantly differ between age groups. These results demonstrate that attentional lapses are associated with transient increase of excitability, and decrease in response propagation and effective connectivity. This pattern is similar to what is observed during sleep, suggesting that lapses reflect a sleep-like phenomenon. These findings could contribute to develop models aimed to predicting and preventing lapses in real life situations.

**Highlights:**

- Cortical excitability increases during attentional lapses
- Brain response propagation is reduced during attentional lapses
- Aging does not affect the differences between normal attention and lapse periods
- Lapses characteristics resemble previous reports made during sleep

## 1. Introduction

Attention is a cognitive process that is required for normal functioning of other cognitive domains. When attention is not focused on the environment, we can experience a detachment that may lead to lapses of attention. Lapses are more prevalent when vigilance decreases, and can contribute to errors (Lim and Dinges, 2008). Their full electrophysiological characterization could therefore contribute to error detection, anticipation and prediction, which is of foremost importance in many disciplines in which they can have catastrophic consequences (e.g. driving, medicine, military, etc.).

The prevalence of attentional lapses is tightly related to the regulation of sleep and wakefulness. During the day, while well rested, they are relatively rare, because sleep need is low and the circadian system helps maintaining wakefulness (Cajochen et al., 1999). If one extends wakefulness during the night beyond habitual sleep time, sleep need further increases while the circadian system promotes sleep, such that lapses become more frequent (Lim and Dinges, 2008; Philip and Åkerstedt, 2006). Invasive recordings in animals have associated lapses with local and transient periods of neuronal silence (OFF-periods), which resemble what happens during sleep (Meisel et al., 2017b; Vyazovskiy et al., 2011). Intracranial recordings in a few epileptic patients showed that neuronal spiking in response to stimulations is attenuated, delayed and lengthened before cognitive lapses (Nir et al., 2017). In addition, slower EEG activity of local field potentials remains relatively high prior to and during lapses (Nir et al., 2017). Similar phenomenon may therefore take place in animals and humans during lapses of attention. However, the extent to which attentional lapses in humans resemble local sleep-like phenomenon during wakefulness, is not fully established. Boundaries of lapses are difficult to define since a lapse often consists in the absence of response to stimulus (Dinges and Powell, 1985). In addition, lapses may alter sensory perception or higher cognitive functions (Chee et al., 2008). In this context, transcranial magnetic stimulation coupled to electroencephalography (TMS-EEG) represents an ideal mean to probe the neural mechanisms underlying lapses. TMS triggers brain responses over a relatively small area of cortex and mimics normal brain functioning, while bypassing sensory inputs and processing (Rosanova et al., 2012). Once EEG is recorded, one can characterize brain responses in terms of shape and propagation (Rosanova et al., 2012).

Cortical excitability reflects the responsiveness and response selectivity of cortical neurons to stimulations and can be probed with TMS-EEG (Rosanova et al., 2012). Its sensitivity to both sleep need and the circadian system has been characterized (Huber et al., 2013; Ly et al., 2016), as well as its changes during sleep (Massimini et al., 2005). Cortical excitability progressively increases with wakefulness extension, with local influence of the circadian system exacerbating the night-time increase (Ly et al., 2016). TMS response propagation varies during prolonged wakefulness. When focusing on the night-time period, when one would be normally asleep, participants with lower response propagation perform worse on a vigilance task, suggesting that a reduction in response spreading at night is associated with worse performance and potentially higher number of lapses (Gaggioni et al., 2018). During slow wave sleep, a further increase of cortical excitability and a limited response propagation is observed (Massimini et al., 2005). Whether similar changes happen during attentional lapses is unknown.

In our increasingly older society, understanding age-related changes in lapses of attention is of timely interest. Modifications in sleep and wakefulness regulation are hallmarks of the aging process (Van Egroo et al., 2019a, 2019b). Sleep becomes “shallower”, more fragmented, and more sensitive to challenges over the adult lifespan (Carrier et al., 2011; Hoel et al., 2016), while the circadian system advances sleep timing and seems to send a weaker sleep and wakefulness promoting signal (Schmidt et al., 2012; Valentinuzzi et al., 1997). However, one suffers less from acute sleep loss in aging, such that lapses are less common in older individuals during sleep deprivation (Duffy et al., 2009). Cortical excitability dynamics during wakefulness extension is also dampened in aging, reflecting both a reduction in strength of both sleep homeostasis and circadian signals (Gaggioni et al., 2019). Whether age-related changes in sleep-wake regulation are reflected in modifications in cortical excitability and brain response propagation during lapses of attention has not been investigated.

Here, we performed a retrospective analysis of TMS-EEG studies to compare TMS-evoked potentials (TEP), as a probe for cortical excitability, and TMS response spatial propagation over the cortex, as a proxy for effective connectivity, during lapses of attention versus normal periods with no-lapse. We collected data recorded at night in healthy younger and older adults, and assessed whether lapses of attention were associated with detectable alteration in cortical excitability and TMS response propagation. Our hypothesis was that cortical excitability and response propagation would, respectively, increase and decrease during lapses, and to a greater extent in younger compared to older individuals.

## 2. Material and methods

Data included in this analysis were retrospectively selected among three different studies, including repeated assessment of cortical excitability using TMS-EEG over the superior frontal gyrus during wakefulness extension protocols (Gaggioni et al., 2019; Ly et al., 2016; Van Egroo et al., 2019a). All studies were approved by the Ethics Committee of the Faculty of Medicine at the University of Liège, Belgium. Participants gave their written informed consent prior to entering the study and received financial compensation.

### 2.1. Participants

Participants’ exclusion criteria were as follows: Body Mass Index (BMI) ≤ 18 and ≥ 29; recent psychiatric history or severe brain trauma; addictions, chronic medication affecting the central nervous system; hypertension; smoking, excessive alcohol (> 14 units/week) or caffeine (> 9 cups/day) consumption; shift work in the past 6 months; transmeridian travel in the past two months; anxiety, as measured by the 21-item self-rated Beck Anxiety Inventory (score ≥ 10) (Beck et al., 1988a); depression, as assessed by the 21-item self-rated Beck Depression Inventory (score ≥ 14) (Beck et al., 1988b). Participants with stable treatment (for > 6 months) for hypertension and/or hypothyroidism were included in the study. Participants with sleep apnea (apnea-hypopnea index ≥ 15/hour) were excluded based on in-lab adaptation and screening night of polysomnography. Older participants with clinical symptoms of cognitive impairment were excluded [Dementia rating scale < 130 (Mattis, 1988) or Mini mental state examination (MMSE) < 27 (Folstein et al., 1975)]. Twelve individuals aged between 18 and 30 years old and 12 individuals aged between 50 and 70 were included in the current analyses (**Table 1**).

**Table 1:**
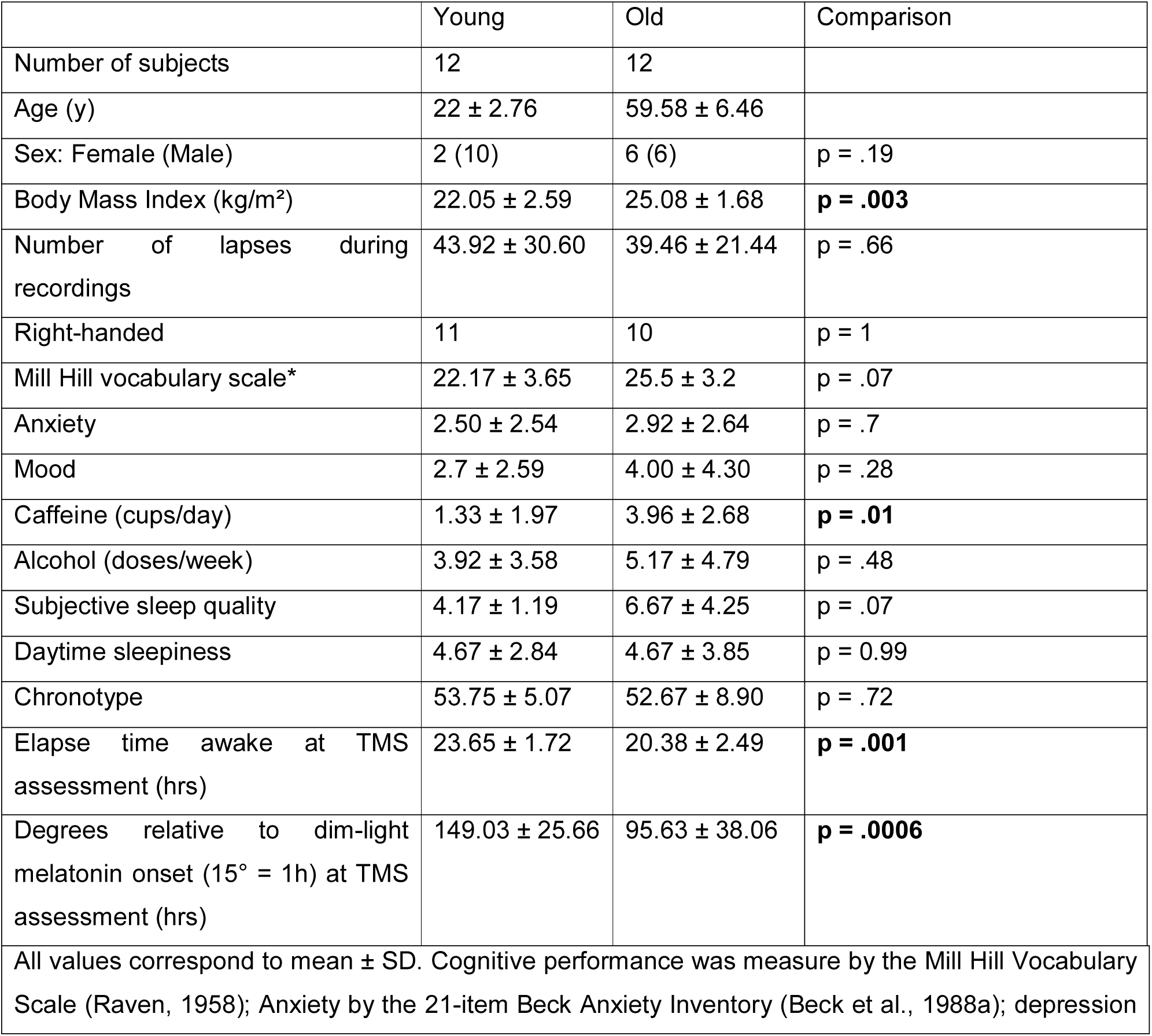
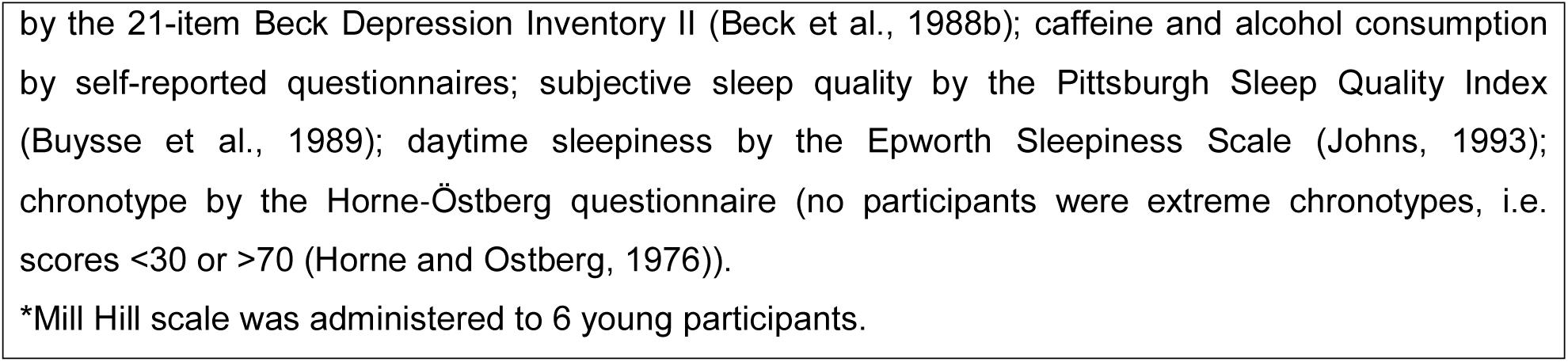
Demographic of the young and older groups

|  | Young | Old | Comparison |
| --- | --- | --- | --- |
| Number of subjects | 12 | 12 |  |
| Age (y) | 22 ± 2.76 | 59.58 ± 6.46 | |
| Sex: Female (Male) | 2 (10) | 6 (6) | p = .19 |
| Body Mass Index (kg/m<sup>2</sup>) | 22.05 ± 2.59 | 25.08 ± 1.68 | **p = .003** |
| Number of lapses during recordings | 43.92 ± 30.60 | 39.46 ± 21.44 | p = .66 |
| Right-handed | 11 | 10 | p = 1 |
| Mill Hill vocabulary scale* | 22.17 ± 3.65 | 25.5 ± 3.2 | p = .07 |
| Anxiety | 2.50 ± 2.54 | 2.92 ± 2.64 | p = .7 |
| Mood | 2.7 ± 2.59 | 4.00 ± 4.30 | p = .28 |
| Caffeine (cups/day) | 1.33 ± 1.97 | 3.96 ± 2.68 | **p = .01** |
| Alcohol (doses/week) | 3.92 ± 3.58 | 5.17 ± 4.79 | p = .48 |
| Subjective sleep quality | 4.17 ± 1.19 | 6.67 ± 4.25 | p = .07 |
| Daytime sleepiness | 4.67 ± 2.84 | 4.67 ± 3.85 | p = 0.99 |
| Chronotype | 53.75 ± 5.07 | 52.67 ± 8.90 | p = .72 |
| Elapse time awake at TMS assessment (hrs) | 23.65 ± 1.72 | 20.38 ± 2.49 | **p = .001** |
| Degrees relative to dim-light melatonin onset (15° = 1h) at TMS assessment (hrs) | 149.03 ± 25.66 | 95.63 ± 38.06 | **p = .0006** |
| All values correspond to mean ± SD. Cognitive performance was measure by the Mill Hill Vocabulary Scale (Raven, 1958); Anxiety by the 21-item Beck Anxiety Inventory (Beck et al., 1988a); depression | | | |

by the 21-item Beck Depression Inventory II (Beck et al., 1988b); caffeine and alcohol consumption by self-reported questionnaires; subjective sleep quality by the Pittsburgh Sleep Quality Index (Buysse et al., 1989); daytime sleepiness by the Epworth Sleepiness Scale (Johns, 1993); chronotype by the Horne-Östberg questionnaire (no participants were extreme chronotypes, i.e. scores <30 or >70 (Horne and Ostberg, 1976)).
\*Mill Hill scale was administered to 6 young participants.

### 2.2. Experimental protocols

Structural MRI was performed on a 3-Tesla MR scanner (MAGNETOM Prisma, Siemens, Germany) through a T1-weighted MPRAGE sequence (*TR = 7*.*92 ms, TE = 2*.*4 ms, TI = 910 ms, FA = 15°, FoV = 256 × 224 × 176 mm^3^, 1 mm isotropic spatial resolution)*. Structural data were used for TMS-neuronavigation and EEG source reconstruction. All participants completed a “pre-test” TMS-EEG session to select the optimal stimulation point over the superior frontal gyrus to avoid muscular or electromagnetic artifacts. Participants were then asked to maintain regular sleep-wake schedules during the 7 days preceding the experiments (+/- 30 min). Compliance was verified using sleep diaries and wrist actigraphy (Actiwatch©, Cambridge Neurotechnology, UK). On the day preceding the experiment, participants arrived at the laboratory 6 to 8 hours before their habitual bedtime and were kept in dim light (< 5 lux) for 5 to 6.5 hours preceding bedtime. They then slept in the laboratory at their habitual sleep and wake times under EEG recording (in darkness, 0 lux). A TMS-compatible EEG cap was placed upon awakening and remained for the whole duration of the protocol. Participants remained awake in dim-light (5 lux) for the 20 (Van Egroo et al., 2019a), 29 (Ly et al., 2016) or 35 (Gaggioni et al., 2019) following wake time, including repeated TMS-EEG assessments. To increase the likelihood of lapses, only night time TMS-EEG recordings were considered in the analyses in each study, following ∼19h (Van Egroo et al., 2019a) and ∼24h (Gaggioni et al., 2019; Ly et al., 2016) of continuous wakefulness, corresponding to 2AM and 7AM for a subject waking up at 7AM (Table 1). To be included in the analyses, the session had to include at least 20 lapses as defined below.

### 2.3. TMS-EEG data acquisition

Stimulation and neuronavigation were achieved with a Navigated Brain Stimulation (NBS) system (Nexstim, Helsinki, Finland) which uses a focal bipulse 8-shape coil equipped with infrared position sensors and a head tracker allowing for coregistration of T1-weighted structural MR images. Recording was done with a 60-channel TMS-compatible EEG amplifier (Eximia, Helsinki, Finland), equipped with a sample-and-hold circuit to provide TMS-artefact-free data from 5 ms post-stimulation (Virtanen et al., 1999). Electrooculogram (EOG) was recorded with two additional bipolar electrodes. EEG signal was band-pass filtered between 0.1 and 500 Hz and sampled at 1450 Hz. Prior to each recording session, electrodes impedance was maintained < 5 kΩ. Auditory EEG potentials evoked by the TMS clicks and sensory stimulation were minimized by playing a continuous pink noise through earphones and applying a thin foam layer between the EEG cap and the TMS coil (Rosanova et al., 2012). Stimulation point was set on the superior frontal gyrus, contralateral hemisphere to subject’s handedness, due to its sensibility to sleep pressure (Huber et al., 2013), the reduced probability to elicit involuntary reaction such as muscular twitches or eye blinks when stimulated, and its direct involvement in the attentive task used (Maquet et al., 2003; Poudel et al., 2013). Each session included around 250 trials, with interstimulus intervals that were randomly set to 1900 to 2200 ms. Participants were continuously monitored by a research staff member during TMS/EEG recording to ensure they would not fall asleep.

### 2.4. Compensatory tracking task

During each TMS/EEG recording, participants were instructed to perform a Compensatory Tracking Task (CTT), a visuomotor vigilance task (Makeig and Jolley, 1996). The task consists in keeping a constantly moving cursor on a central circular target, using a trackball device. Performance is measured as the distance, in pixels, between the cursor and the target. Transitory lapses of attention immediately result in temporary increases of the target-cursor distance. A lapse was defined as a time when the cursor was located outside of a central 200 by 200 pixels box surrounding the target and >500 ms from the last trackball movement. TMS-evoked responses occurring during and <1s around a lapse were considered as acquired during a lapse. CTT was preferred to other classic lapse measures, such as the psychomotor vigilance task (PVT) (Dinges and Powell, 1985), because it does not need the burst-like muscular activity time-locked to the TEP but rather requires continuous smooth and limited movement of a single finger.

### 2.5. TMS-EEG data processing

Data were visualized and processed in MATLAB 2015 (The Mathworks Inc, Natick, MA). Data were visually inspected to reject trials with magnetic artifacts and eye movements. Noisy and artifacted channels were rejected. Data were highpass-filtered at 1 Hz, then downsampled to 1000 Hz and finally lowpass-filtered at 80 Hz. Individual trials were then epoched between -100 and 300 ms post TMS. Baseline correction between -100 and -1.5 ms was applied before averaging across trials, using robust averaging method (Leonowicz et al., 2005) (**Figure 1A**).. Then, the electrode closest to the TMS-EEG target in the stimulation hemisphere was chosen to extract excitability. Excitability of the cortex was inferred based on the first component of the averaged TEP (0-30 ms; **Figure 1B**). Amplitude and maximal slope (referred to as slope) were the main parameters extracted to define cortical excitability, together with the latencies to the first negative and positive peaks of the TEP.

**Figure 1:**
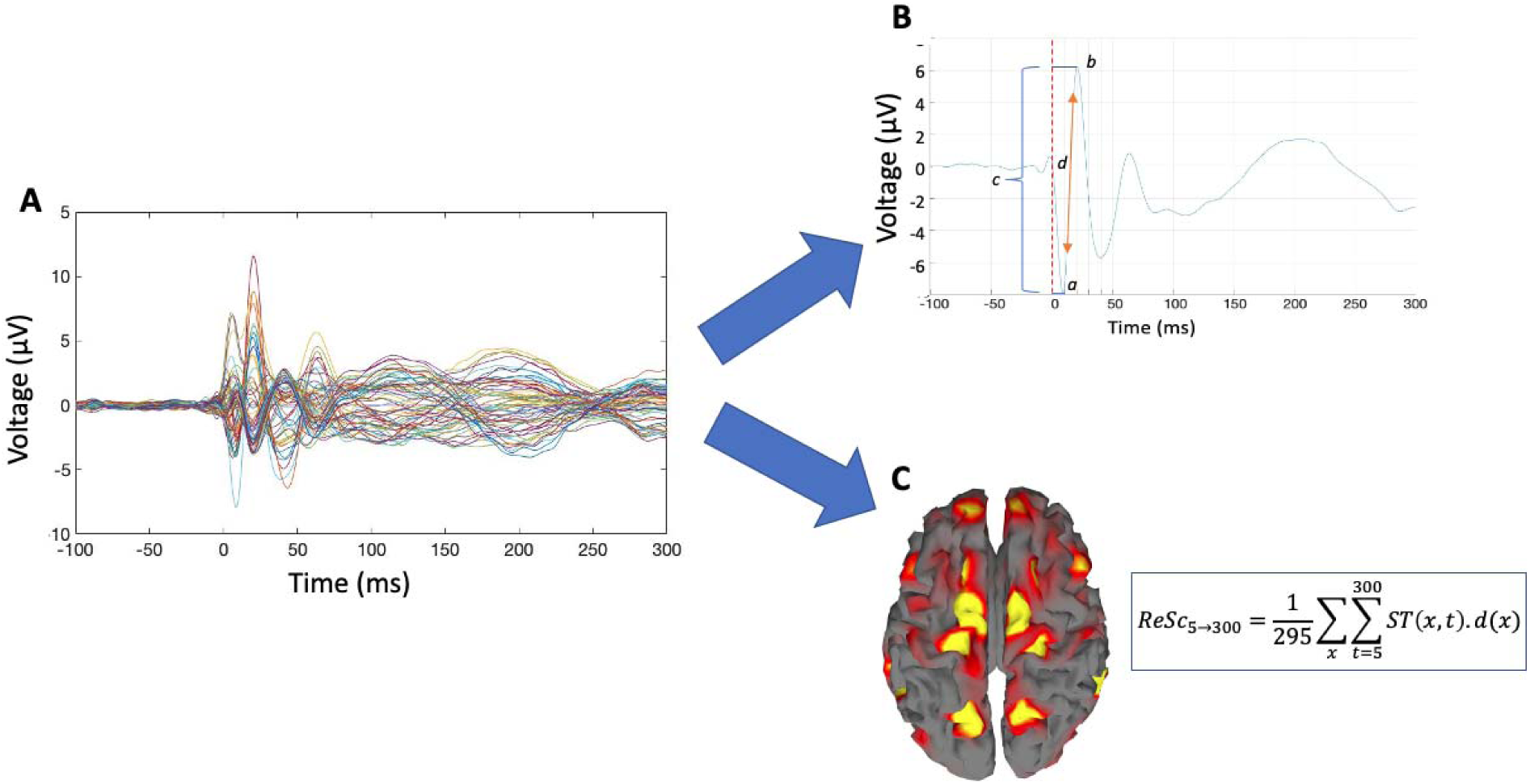
Overview of EEG data processing. **A**. Butterfly plot of the average TMS responses over all 60 channels in a representative subject. **B**. Average TMS response of the electrode closest to the stimulation point, from which the following are extracted *a)* Latency of the negative peak *b)* Latency of the positive peak *c)* Peak to Peak amplitude *d)* Steepest slope, i.e. the tangent at the inflection point. **C**. Response scattering (ReSc) was computed following EEG source reconstruction and binarization of the significant and non-significant sources in time and space. ReSc is computed from this spatiotemporal binary matrix (ST) from 5 ms post-TMS to 300 ms post-TMS (*t*) as the sum of geodesic distance (*d*) between significant sources (*x*) and the TMS target.

Source reconstruction followed the procedure previously described (Gaggioni et al., 2018). Briefly, the averaged TMS-evoked EEG response from 0 to 300 ms post-TMS pulse on all available channels was used to obtain a spatio-temporal matrix of significant cortical sources. Sensor and fiducial positions were used for realistic head model with Boundary Element Method (BEM) constructed based on individual MRI, sensor and fiducial positions, to perform all the analyses within the individual subject space. The inverse solution was based on the “Multiple Sparse Prior” (MSP) method with 5124 dipoles to model the propagation of significant current in the brain induced by the stimulation. Response Scattering (ReSc) consist in the sum of the geodesic distance between significant sources and the TMS target, averaged over the entire 5– 300 ms period post-TMS **Figure 1C**).

### 2.6. Statistics

Statistical analyses were performed with SAS version 9.4 (SAS, Institute, Cary, NC, USA). Outlier threshold was set at 3 standard deviations from the mean, but no outlier values were detected. TEP amplitude, slope and latencies as well as ReSc constituted dependent variables of separate Generalized Linear Mixed Models (GLMM; PROC GLIMMIX SAS procedure), considering subject as random factor and response type (lapse, normal) as repeated measures with autoregressive correlation type 1 (ar(1)). Dependent variable distributions were estimated using the using “allfitdist” function in MATLAB (developed by Mike Sheppard) and set accordingly in each GLMM. Models included also included the following covariates: age group, study, sex, BMI and TMS parameters (mean generated electric field at the hotspot and electrode of interest distance from hotspot). Since the session considered in each studies were acquired at different circadian phase, an interaction between study and response type was included in the model to take into account potential non-linear changes in response type effect with circadian phase. Similarly, we included a possible interaction between age groups and lapses for possible age-specific modulation of the measured parameters during lapses. Partial effect sizes of the significant effects were calculated based on semi-partial R-squared 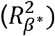 computation for GLMM according to the literature (Jaeger et al., 2017). We set the significance threshold to p-value = 0.01, accounting for Bonferroni correction for testing five models.

## 3. Results

As indicated in **Table 1**, except for BMI and caffeine consumption where older individuals had higher values, age groups did not differ for any demographic factor such as sex of the participants and number of lapses per sessions. The difference between age-groups in time elapsed awake and in degrees from DLMO is linked with different timing of the session considered in each studies; it is taken into account in GLMM by including study as covariate.

When assessing differences between electrophysiological measures between lapses and no-lapses, we first considered slope of TEP as the most typical assessment of cortical excitability (Huber et al., 2013; Ly et al., 2016). GLMM including sex, age, BMI, study and TMS parameters as covariates indicated that response type (Lapse vs No-Lapse) significantly affected TEP slope, with higher slope for lapse compared to no-lapse periods (p = .009) representing a large effect size 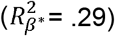 (**Figure 2A,E; Table 2**). Similarly, TEP amplitude showed an effect of response type (p = .0003) with higher amplitude for lapse vs. normal attention representing a large effect size 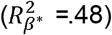) (**Figure 2B,E; Table 2**). We further found that latencies of the first TEP negative and positive peaks were, respectively, shorter and longer for lapses vs. no lapses (negative peak: p = .0007; large effect size, 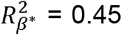 positive peak: p = .008, large effect size: 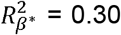 (**Figure 2C-E; Table 2**).

**Table 2:**
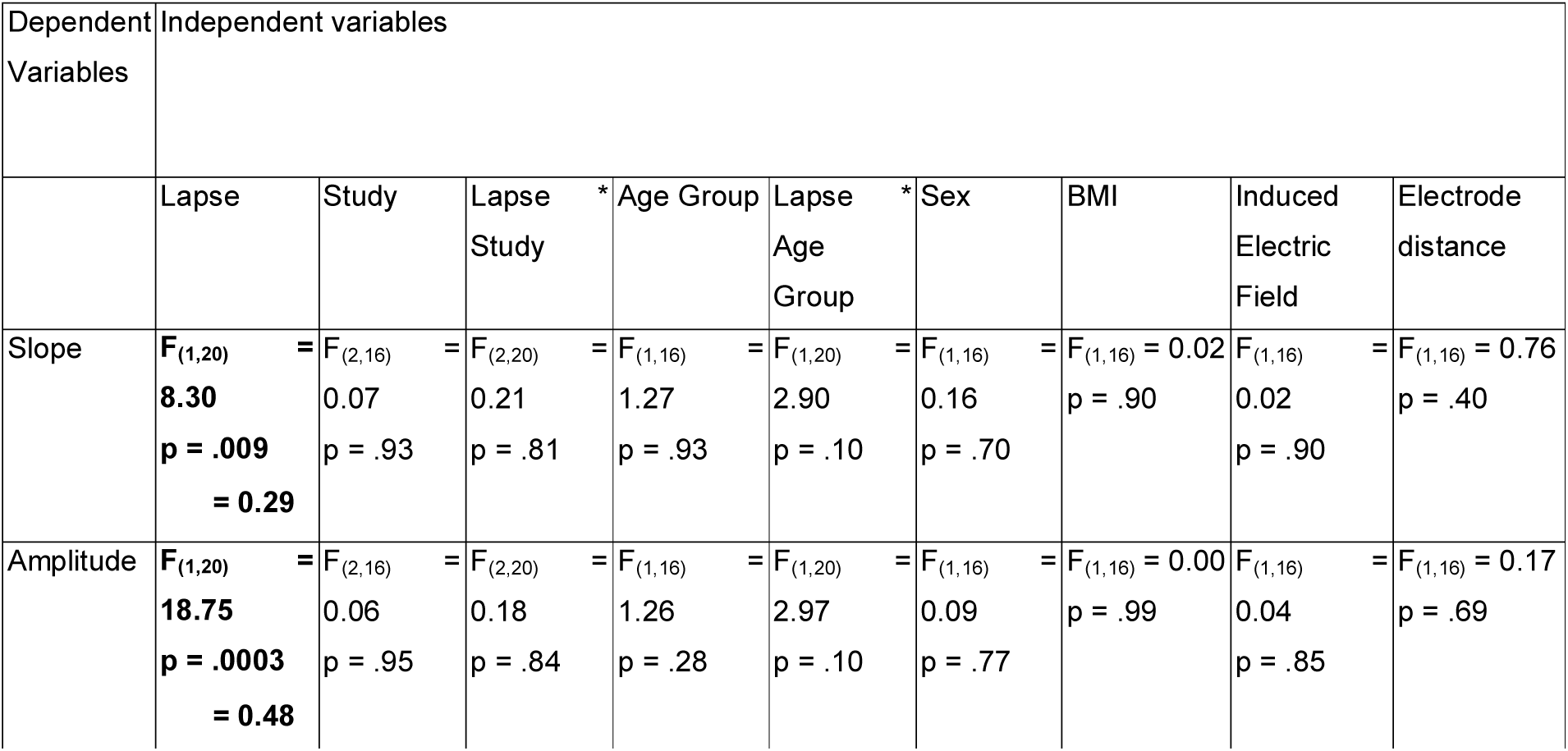
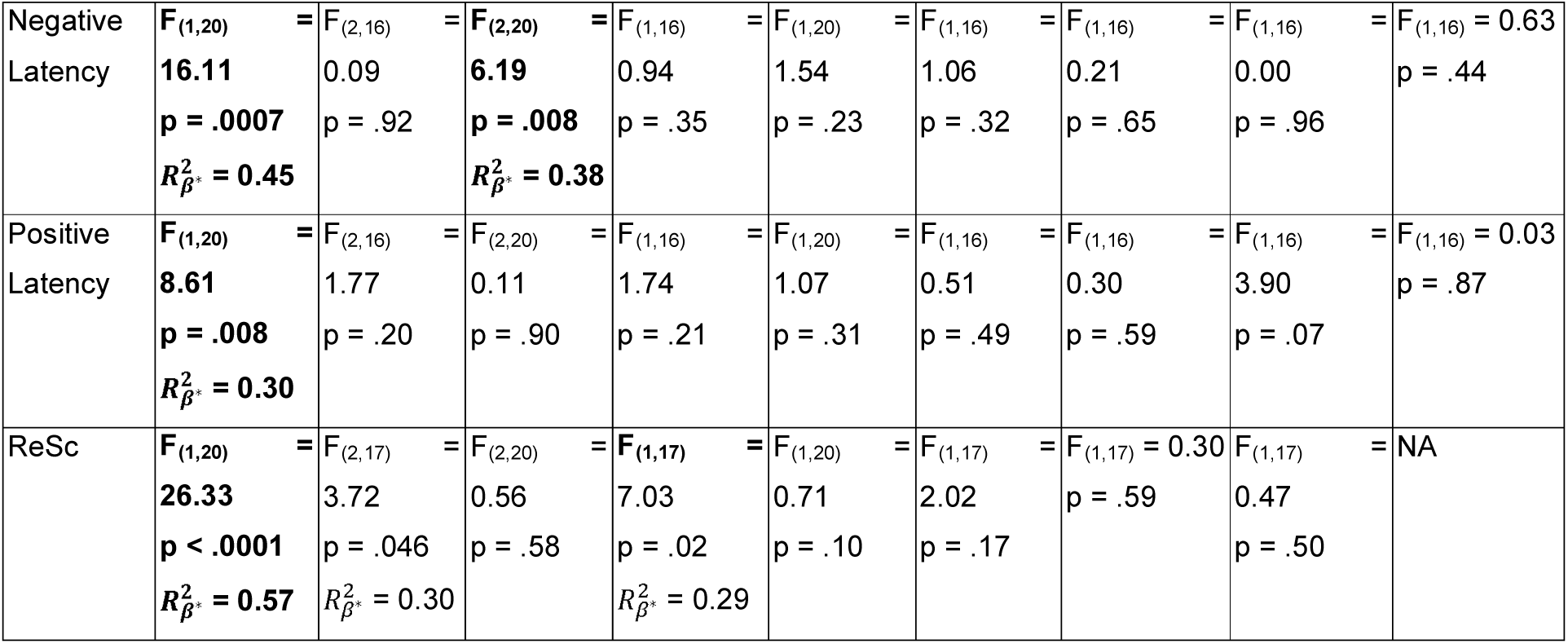
GLMM outcomes for the four excitability measurements and response scattering (ReSc). Significant results (following multiple testing correction; p = 0.01) are in bold. When a variable is significant, the partial effect size is reported.

**Figure 2:**
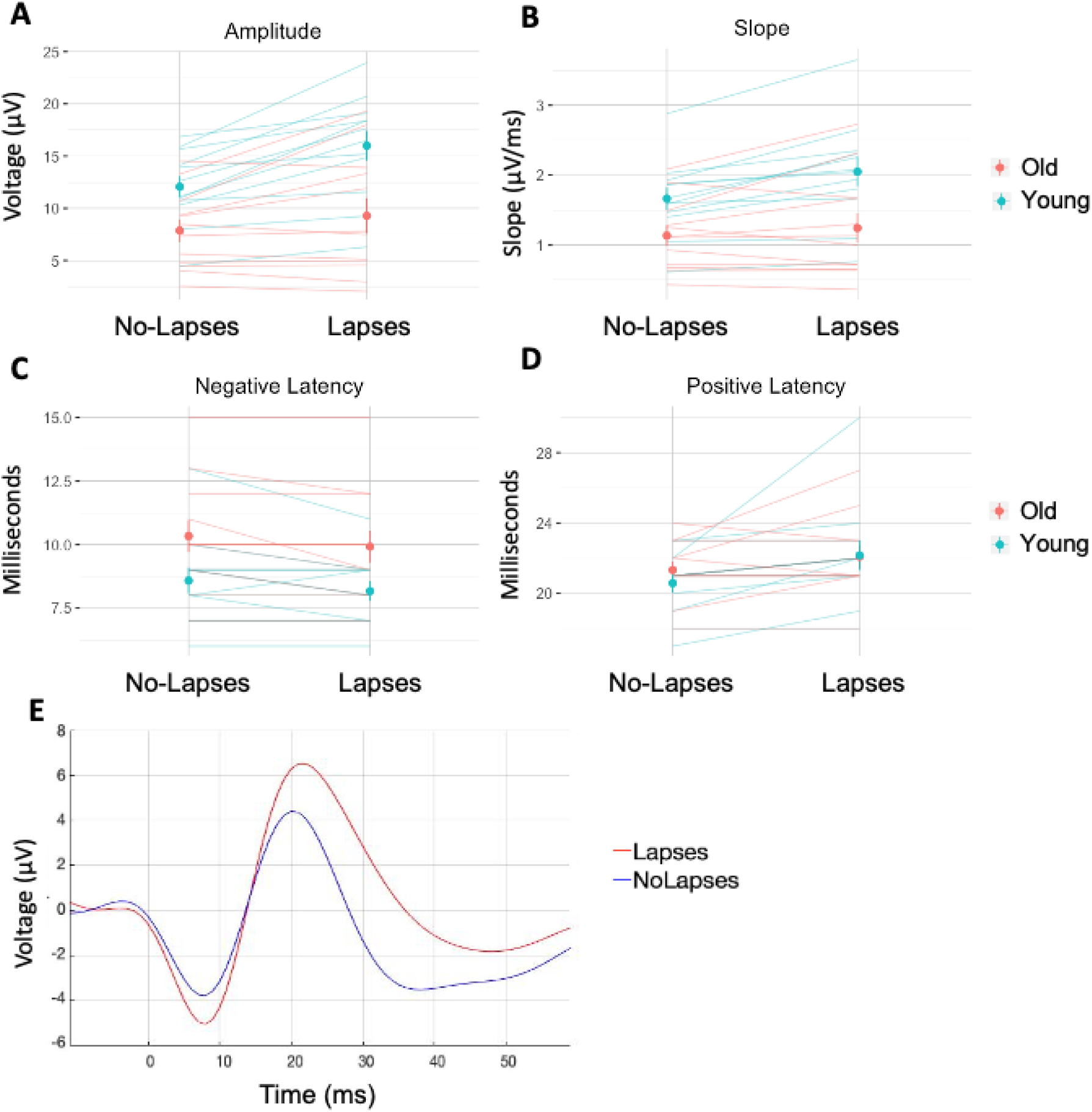
Amplitude (A), slope (B), negative (C) and positive (D) peak latencies during lapse and no-lapse periods in both age groups (Red = Older, Blue = Young). Individual lines represent single- subject values. Dots are the group means and the vertical bars represent the standard error. (E) TEP during lapse and no-lapse in a representative subject.

Importantly, age - as well as all the other covariates of the models - was not significantly associated with any of the 4 TEP parameters, suggesting that differences between lapses vs. no- lapse did not differ across age group (**Table 2**). We obtained a significant interaction between study and lapses for the latency of the first negative peak. Post-hoc analyses point to a larger difference between lapse and no-lapse in one of the studies (Ly et al., 2016) compared to the other studies for latency to negative peak (p = .008), but, overall, all studies show similar direction in the difference between lapse and no-lapse (i.e. lapse < no-lapse).

We then turned to TMS Response Scattering (ReSc) to assess whether TMS response propagation would differ between lapse and no-lapse periods. GLMM yielded a significantly reduced ReSc during lapses as compared to no-lapse (p < .0001) with a large effect size (= 0.57) (**Figure 3; Table 2**). A significant effect of study and age were also detected, with older individuals showing reduced ReSc (p = .02, = 0.29), but these effects did not reach corrected statistical significance. To assess whether the local cortical excitability differences between lapses and no-lapse reported above could have contributed to differences in ReSc, we added TEP slope in the model. TEP slope was not associated to ReSc (F_(1, 19.16)_ = 0.17, p = .68), while the inclusion of TEP slope only marginally affected the difference in ReSc between lapses and no-lapse (F_(1, 24.47)_ = 32.08, p < .0001).

**Figure 3:**
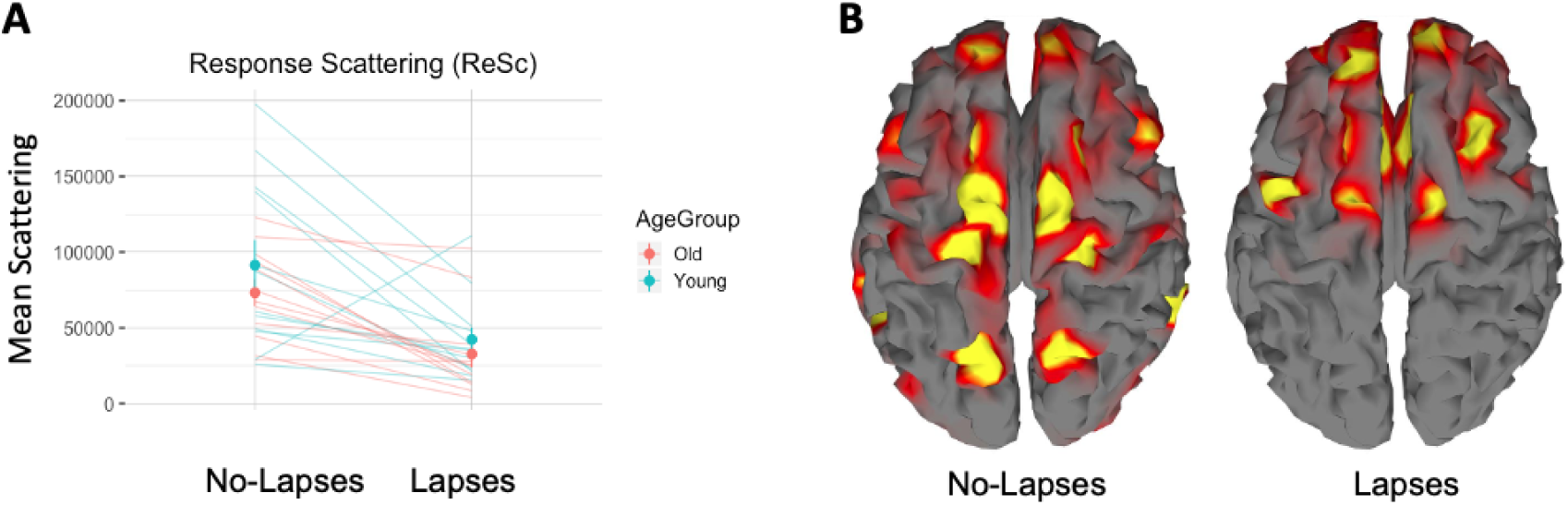
Response Scattering (ReSc) during lapse and no-lapse periods. A) ReSc during lapses and no-lapses in both age groups (Red = Old, Blue = Young). Individual lines represent single-subject values. Dots are the group means and the bars represent the standard error. B. Significant sources reconstructed at xxms post-TMS in a representative subject, showing that less numerous and less distant sources are detected during lapses.

## 4. Discussion

Lapses of attention are common during night-time wakefulness and contribute to errors. Previous invasive animal and human studies have related lapses to local sleep-like periods during wakefulness over limited portion of the brain. Here, we used TMS-EEG to measure cortical excitability and brain response propagation during lapse and no-lapse periods while performing a continuous vigilance/sustained attention task. We report that, during attentional lapses, the first component of TEP shows a significant increase in slope, amplitude, and latency to its positive peak together with a shorter negative peak latency. In line with our hypothesis, these results demonstrate that cortical excitability is increased during lapses of attention. We further show that TMS response propagation, as indexed by ReSc, was significantly lower during lapses. This indicates that TMS response remains more local during lapses. Taken together, these findings are reminiscent of previous observations made during sleep (Massimini et al., 2005), supporting that attentional lapses resemble a transient “sleep-like state”. Finally, and contrary to our expectation, we find that the difference in cortical excitability during lapse and no-lapse periods does not differ between age groups, while ReSc may be further reduced during lapses in older individuals compared to young individuals, although the difference did not reach corrected statistical significance.

Animal data showed that local neuronal silent periods are more prevalent with increasing wakefulness duration and sleep deprivation, similarly to neuronal off-periods, or down-states, during sleep (Vyazovskiy et al., 2011). During sleep, these off-periods contribute to neuronal firing synchrony to generate the typical EEG slow waves, while during wakefulness off-periods are associated to slower local field potential variations and reduced performance. Similarly, investigation in epileptic human patients indicates that attentional lapses during normal wakefulness are preceded and concomitant to attenuated, delayed and lengthened spiking of individual neurons in the medial temporal lobe (Nir et al., 2017). These effects appeared exacerbated following sleep deprivation, although based on a few patients only. The increased TEP slope we observe during lapses likely arises in part from neurons being more synchronously recruited, potentially because of more prevalent off-periods. Furthermore, larger TEP amplitude during lapses is also likely due to more neurons contributing to the response, maybe because of transient changes in neuronal membrane potentials, which could be related to temporary fluctuations in neuromodulator levels, such as acetylcholine (Parikh et al., 2007) or norepinephrine (Mittner et al., 2016). In other words, some neurons would be responding to the TMS pulse, which mimics normal stimulus brain processing, when they would not outside a lapse, resulting in altered cortical neuron response selectivity.

The reduced response propagation we detected during lapses further implies an existing alteration in long-range cortical processing. During lapses, less areas and less distant areas are recruited in response to a stimulation, whether or not we take into account the initial increase in cortical excitability. This could again be related to more prevalent silent neuronal periods. Animal data demonstrated that the intrusion of an off-period compromises signal propagation in time during sleep deprivation (Meisel et al., 2017b). A similar reduction of spontaneous scalp EEG signal propagation in time at quiet rest was observed in healthy human beings, potentially due to silent periods (Meisel et al., 2017a). Here, we extend these findings to the spatial propagation of brain responses triggered by external stimulations. Overall, our findings imply that higher cortical processing is altered during lapses, i.e. outside sensory cortices and independent of potential changes in their initial responses, since TMS triggers direct responses by-passing sensory systems. This alteration is likely to contribute to slower behavioral responses, reduced performance, and/or absence of response.

Increased cortical excitability and reduced response propagation are observed during healthy human sleep (Massimini et al., 2005). We report similar changes during lapses as compared to normal attention periods. Compared with prior reports, we extend the similarities between lapse and local sleep-like brain activity to healthy individuals and to non-invasive recording of neurons to direct cortical stimulation. Intracranial recording in patients showed that neurons have an increased tendency to fall into an off-period in response to a direct stimulation (Pigorini et al., 2015). If this tendency was increasingly present when wakefulness is extended beyond its normal duration, this could mean that rather than depending on overall more prevalent silent periods or off-periods during sleep deprivation, increased excitability and reduced response propagation during lapses could result, in part, from a large number of neurons entering off-periods in response to TMS. This would also result in higher cortical excitability and more local responses, and would affect sensory processing up to higher cortical areas. We are not in a position to assess the validity of this assumption. We also cannot assess a potential contribution of subcortical structures, such as the thalamus, and thalamo-cortical loops, to the altered propagation we detect.

Contrary to our expectations, our results suggest that, despite the large impact of aging on sleep and wakefulness regulation (Schmidt et al., 2012; Valentinuzzi et al., 1997), age was not associated with strong modifications in the electrophysiological characteristics of lapses. The reduced prevalence of lapses during acute sleep deprivation in older individuals, that is associated with more stable sleepiness and EEG spectral composition (Landolt et al., 1996; Münch et al., 2004), does not appear to be underlined by changes in the phenomenology of lapses *per se*. This statement would need to be confirmed in a larger sample, particularly, for response propagation during lapses.

EEG recordings of TMS responses can be used to study cortico-cortical interactions from a causal perspective. In that respect, ReSc constitutes an index of effective connectivity. The reduction in TMS response propagation during sleep was interpreted as a reduced effective connectivity (Massimini et al., 2005). This finding also led to the conclusion that the fading of consciousness during slow wave sleep was associated with effective connectivity. The implication of our findings for consciousness is however unclear. Our participants did not sleep during the TMS-EEG recordings – even micro-sleep –, as they were closely monitored, and they were obviously conscious. Although one can arguably postulate that their consciousness was different during lapse vs. no-lapse periods and that participant’s brain may experience some local sleep-like phenomenon during lapse (Andrillon et al., 2019), it is difficult to argue that, during lapses, they were experiencing an intermediate level of consciousness between fully rested daytime wakefulness and unconscious sleep. Qualitative comparisons with previous observation during sleep (Massimini et al., 2005) may suggest that TMS responses remain more local during slow wave sleep than during nighttime lapses. Yet, this cannot be certified through quantitative comparisons due to differences in data acquisitions and analyses. We therefore cannot assess whether all our conscious participants were presenting a higher ReSc level than a given threshold that would delineate the transition to unconsciousness, as it has been done using other indices (Casali et al., 2013).

In addition, whether cortical excitability and brain response propagation (and effective connectivity) are qualitatively or quantitatively similarly affected by attentional lapses after sleep deprivation or by those, more sporadic, detected during normal rested wakefulness, remains to be assessed. Night-time/sleep deprivation lapses are likely to favor sleep onset, while daytime lapses are less likely to do so, such that one can expect brain activity differences. Moreover, since we only stimulated the frontal cortex, which shows the largest increase in sleep slow wave prevalence following sleep loss (Cajochen et al., 2001), we expect regional differences in cortical excitability changes associated to lapses. Given that sleep slow wave power shows a relative increase over the entire brain following sleep loss, we expect a relative increase in excitability during lapses whatever the stimulated brain region (Maric et al., 2017), but quantifying these local variations will require further investigations. Likewise, since ReSc is a global brain measure, we consider that our findings reflect a whole-brain reduction in effective connectivity. This does not preclude however the origin of the initial brain response to affect its propagation. Finally, beyond their similarities with sleep, lapses of attention, at night and during the day, could share similar brain activity features with mind wandering or mind blanking (Kawagoe et al., 2019) or may be one of the sheer brain hallmarks of “errors”.

In conclusion, we report that during nighttime attentional lapses, cortex excitability is higher while brain responses remain more local, similar to what happens during sleep. The relevance of these findings is not limited to the theoretical understanding of sleep and wakefulness regulation but may help online detection of lapses and interventions to prevent errors (e.g. through closed-loop electrophysiological stimulation).

## Data Availability

The data that support the findings of this study are available from the corresponding authors upon reasonable request.

## Acknowledgements

We thank E. Balteau, C. Bastin, G. Besson, S. Chellappa, F. Collette, C. Degueldre, C. Hagelstein, B. Herbillon, P. Hawotte, E. Lambot, B. Lauricella, A. Luxen, J.Q.L. Ly, D. Marzoli, V. Muto, P. Maquet, X. Pépin, C. Phillips, E. Salmon, C. Schmidt, and E. Tezel for their help in the different studies.

MVE was and GV is supported by the Fonds de la Recherche Scientifique - FNRS-Belgium. PC and JN were supported by ULiège. GG was supported by Wallonia Brussels International (WBI) and Fonds Léon Frédéricq. The study was supported by the Wallonia-Brussels Federation (Actions de Recherche Concertées - ARC - 09/14-03 & 17/27-09), Walloon Excellence in Life Sciences and Biotechnology Grant (WELBIO-CR-2010-06E), FNRS-Belgium (FRS-FNRS, F.4513.17 & 3.4516.11), University of Liège (ULiège), Fondation Simone et Pierre Clerdent, Fonds Léon Frédéricq and European Regional Development Fund (Radiomed project).

## Author Contribution

PC, MVE, JN, DC, GG and GV collected and preprocessed the data. Data reanalysis: PC. Manuscript drafting: PC, GV. Manuscript editing: All.

